# Use of *Andrographis paniculata* (Burm.f.) Wall. ex Nees and risk of pneumonia in hospitalised patients with mild COVID-19: a retrospective cohort study

**DOI:** 10.1101/2022.01.01.22268609

**Authors:** Jeeranan Tanwettiyanont, Napacha Piriyachananusorn, Lilit Sangsoi, Benjawan Boonsong, Chamlong Sunpapoa, Patcharawan Tanamatayarat, Nat Na-Ek, Sukrit Kanchanasurakit

**Affiliations:** Division of Clinical Pharmacy, Department of Pharmaceutical Care, School of Pharmaceutical Sciences, University of Phayao, Phayao, 56000 Thailand; Division of Pharmaceutical Care, Department of Pharmacy, Phrae Hospital, Phrae, 54000 Thailand; Division of Internal Medicine, Department of Nurse, Phrae Hospital, Phrae, 54000 Thailand; Division of Pharmacy and Technology, Department of Pharmaceutical Care, School of Pharmaceutical Sciences, University of Phayao, Phayao, 56000 Thailand; Unit of Excellence Technologies for Natural Products and Herbs, School of Pharmaceutical Sciences, University of Phayao, Phayao, 56000 Thailand; Pharmacoepidemiology, Social and Administrative Pharmacy (P-SAP) research unit, School of Pharmaceutical Sciences, University of Phayao, Phayao, 56000 Thailand; Division of Social and Administration Pharmacy, Department of Pharmaceutical Care, School of Pharmaceutical Sciences, University of Phayao, Phayao, 56000 Thailand; Center of Health Outcomes Research and Therapeutic Safety (Cohorts), School of Pharmaceutical Sciences, University of Phayao, Phayao, 56000 Thailand; Unit of Excellence on Clinical Outcomes Research and IntegratioN (UNICORN), School of Pharmaceutical Sciences, University of Phayao, Phayao, 56000 Thailand; Unit of Excellence on Herbal Medicine, School of Pharmaceutical Sciences, University of Phayao, Phayao, 56000 Thailand

**Author notes:** **Correspondences:** Dr. Nat Na-Ek, PharmD MSc PhD, Assistant Professor Sukrit Kanchanasurakit, PharmD.

**Keywords:** COVID-19, *Andrographis paniculata*, andrographolide, pneumonia, hospitalisation

## Abstract

**Background:** *Andrographis paniculata* (Burm.f.) Wall. ex Nees (AP) has been widely used in Thailand to treat mild COVID-19 infections since early 2020; however, supporting evidence is scarce and ambiguous. Thus, this study aimed to examine whether the use of AP is associated with a decreased risk of pneumonia in hospitalised mild COVID-19 patients.

**Methods:** We collected data between March 2020 and August 2021 from COVID-19 patients admitted to one hospital in Thailand. Patients whose infection was confirmed by real-time polymerase chain reaction, had normal chest radiography and did not receive favipiravir at admission were included and categorised as either AP (deriving from a dried and ground aerial part of the plant), given as capsules with a total daily dose of 180 mg of andrographolide for five days or standard of care. They were followed for pneumonia confirmed by chest radiography. Multiple logistic regression was used for the analysis controlling for age, sex, diabetes, hypertension, statin use, and antihypertensive drug use.

**Results:** A total of 605 out of 1,054 patients (mostly unvaccinated) were included in the analysis. Of these, 59 patients (9.8%) developed pneumonia during the median follow-up of 7 days. The incidence rates of pneumonia were 13.93 (95% CI 10.09, 19.23) and 12.47 (95% CI 8.21, 18.94) per 1,000 person-days in the AP and standard of care groups, respectively. Compared to the standard of care group, the odds ratios of having pneumonia in the AP group were 1.24 (95% CI 0.71, 2.16; unadjusted model) and 1.42 (95% CI 0.79, 2.55; fully adjusted model). All sensitivity analyses were consistent with the main results.

**Conclusions:** The use of AP was not significantly associated with a decreased risk of pneumonia in mild COVID-19 patients. While waiting for insights from ongoing trials, AP’s use in COVID-19 should be done with caution.

## Introduction

*Andrographis paniculata* (Burm.f.) Wall. ex Nees (AP), also known as ‘Fa Thalai Chon’, has been widely used in Thailand for treating upper respiratory tract infections and noninfectious diarrhoea for decades.(1) The main phytochemical constituent of the aerial parts of AP is a diterpenoid lactone compound called ‘andrographolide’, which has shown antiviral and immunomodulatory properties in preclinical and clinical studies.(2–4) Recently, an *in silico* study showed the potential effect of andrographolide on SARS-CoV-2, as the compound can bind and inhibit the viral protease enzyme and viral spike glycoprotein.(4–6) Moreover, *in vivo* and *in vitro* studies consistently supported the effect of AP extract on COVID-19 infections.(7,8)

In addition to preclinical studies, two small clinical trials using a high dose of AP extract to treat mild COVID-19 infections have shown its efficacy in terms of reducing COVID-19 symptoms (e.g., fever, sore throat, rhinorrhoea, cough, headache, anosmia, myalgia, and diarrhoea) (9) and C-reactive protein (CRP) levels.(10) However, its efficacy on important clinical outcomes, especially pneumonia, is unclear.(10) Currently, five ongoing trials are investigating the efficacy of AP in terms of pneumonia for treating mild COVID-19 cases (Table S2, supplementary appendix). Additionally, one trial of Xiyanping injection (andrographolide derivatives) showed promising results; however, the efficacy of oral administration cannot be extrapolated.(11)

Although AP’s efficacy on the risk of pneumonia in COVID-19 is still ambiguous,(10) its widespread use has been encouraged. This is due to the situation in which Thailand experienced a shortage of favipiravir and COVID-19 vaccines at the start of a new pandemic wave in early 2020. Therefore, a pharmacovigilance study is necessary to support the decision of clinicians and policymakers on whether AP’s use in COVID-19 should be further supported.

In this study, we primarily aimed to use real-world data to investigate whether the use of AP was associated with better clinical outcomes in hospitalised mild COVID-19 patients. We also examined the course of COVID-19 and the incidence of pneumonia due to COVID-19 in a country-specific context. Our ultimate goal is to make the best use of available data to inform the public and improve patient care.

## Materials and Methods

The report of this study followed the Strengthening the Reporting of Observational Studies in Epidemiology (STROBE) guidance for reporting cohort studies (Table S1).(12)

### Design, setting, and study population

This is a single-centre retrospective cohort study in which the data were collected from medical records of patients diagnosed with COVID-19 infection. We used the 10^th^ revision of the International Classification of Diseases (ICD-10) code U07.1 to identify potential participants from 1^st^ March 2020 to 31^st^ August 2021. The ethical committee for clinical research of Phrae Hospital approved this study (no. 70/2564). The setting of our study is Phrae Hospital, a 500-bed secondary hospital located in northern Thailand. Eligible participants were at least 18 years old and diagnosed with COVID-19 infection by real-time polymerase chain reaction (RT–PCR). According to the definition of mild COVID-19 used in previous work,(11,13) we included only patients who had normal chest radiography by the time of admission. In contrast, individuals who did not have chest radiography results, received favipiravir, or received systemic corticosteroids on the first admission date were excluded. In addition, we also excluded those who took AP prior to admission, had a history of allergy to AP, had elevated liver enzymes, or were pregnant or breastfeeding from the analysis. Since the preliminary data suggested that AP’s efficacy was shown if it was given to patients as soon as they were diagnosed, we additionally excluded patients who received AP after five days of admission from our analysis.(14)

### Exposure

Included participants who received AP within five days of admission in addition to supportive treatment were categorised as an exposed group. AP was prepared as a capsule of 500 mg of a dried and ground aerial part of the plant. Each 500-mg capsule contains an andrographolide content of approximately 4% w/w (20 mg/capsule). According to a previous trial,(10) the AP product was given three capsules thrice daily after a meal to reach a total dose of andrographolide 180 mg/day for five days. Song Hospital, Phrae, Thailand, produced the AP product used in this setting. The quality of the AP product was tested and certified by the Medicinal Plant Research Institute and the regional Medical Sciences Centre, Chiang Rai, Thailand (Supplementary appendix). Supportive treatment, including antipyretics, mucolytics, expectorants, antihistamines, oral rehydration salts, and anxiolytics, was given to patients who did not receive AP (unexposed group).

### Outcomes

The primary outcome was developing pneumonia based on chest radiography during hospital admission. The diagnosis of pneumonia was based on chest X-rays (CXR) of category four or above according to the Modified Rama-Co-RADS criteria (Supplementary appendix) made by infectious disease physicians or radiologists. All patients were followed until being discharged alive or died.

In addition, we analysed the association between receiving AP and a secondary outcome, which was a composite of receiving favipiravir, systemic corticosteroids, or ventilator support; having oxygen saturation drop along with worsening signs and symptoms; or presenting regressive CXR findings (i.e., category three or above) after admission. The CXR results, all clinical data, and relevant medications were collected from electronic medical records.

### Covariates

We collected all covariates for the admission date from medical records. These covariates included age, sex, weight, height, comorbidity, current medications, and laboratory parameters. According to our proposed directed acyclic graphs (DAGs, Figure S1), Table S5 and previous works,(15,16) age, body mass index, hypertension, type 2 diabetes (T2DM), ACEIs/ARBs, statins, and COVID-19 severity were considered confounders. Admittedly, during the data collection period, there were only two patients who previously received a COVID-19 vaccine. Consequently, we did not include vaccination profiles in the analysis.

### Statistical methods

In this study, we included all eligible patients in the analysis. Therefore, sample size calculation was unnecessary, and we calculated statistical power afterwards. Descriptive and inferential statistics were used to compare participants’ characteristics at hospital admission according to their exposed groups. In addition to the calculated incidence rate of pneumonia according to exposed groups, a Kaplan–Meier plot for the probability of a pneumonia-free event between groups was also created and statistically compared using a log-rank test.

The main analysis was performed using a multivariable logistic regression based on a complete-case approach. The justification for using a logistic model is that each participant had a relatively similar follow-up time and the incidence of pneumonia in COVID-19 patients was approximately 10% from a previous trial.(10) To investigate the association between receiving AP and incident pneumonia, we performed serial adjustment as follows: 1) unadjusted model, 2) age-adjusted model, and 3) full adjustment (i.e., adjusting for age, hypertension, T2DM, ACEIs/ARBs, and statins). Regarding BMI, we further performed multiple imputations by chained equations (MICE) to impute missing values. BMI was then included in a model as part of a sensitivity analysis since missing BMI values were unlikely to be under a missing at random (MAR) mechanism and using MICE might bias the results. We performed 100 imputations, and the results were combined using Rubin’s rule.

For the sensitivity analysis, we analysed the data using Cox’s proportional hazards model stratified by diabetes. The Schoenfeld residuals test and log-minus-log plots were used to test the proportional hazards assumption. Moreover, the severity of COVID-19 was conditioned by restricting the analysis to a mild case only. Furthermore, we performed subgroup analyses according to sex, age group (i.e., <60, ≥60), hypertension, T2DM, ACEIs/ARBs, and statin use. Last, to minimise a cohort effect due to differences in admission period (Figure S3) and the effect of receiving COVID-19 vaccination, we excluded individuals admitted before the 1^st^ of July 2021 and two participants who received at least one shot of COVID-19 vaccine prior to admission then re-analysed accordingly.

All analyses were performed using STATA version 16.1 MP (StataCorp LLC, College Station, Texas) and R version 3.3 with a two-sided alpha error of 5%. As we did not adjust for multiplicity, findings of the secondary outcome, sensitivity analyses, and subgroup analyses should be used for exploratory purposes only.

## Results

Among 1,054 COVID-19 patients admitted to the hospital between March 2020 and August 2021, 605 were included in the final analysis (Figure 1). Of these, 351 individuals (58%) received AP within five days of admission. Regarding the characteristics of the included participants at hospital admission (Table 1), the majority of the participants were male (50.4%), with a mean age of 35.41 years old and a mean BMI of 24.2 kg/m^2^. Only a small proportion of individuals had hypertension (7.3%), T2DM (2.2%), and cardiovascular disease (0.8%). In addition, 3.8% and 2.6% of the patients received ACEIs/ARBs and statins, respectively. Comparing between groups, most of the characteristics were relatively similar, except for alkaline phosphatase (ALP) levels, as the levels in the AP group were slightly higher than those in the standard of care group. However, all laboratory parameters were within the normal range (Table 1).

**Figure 1.**
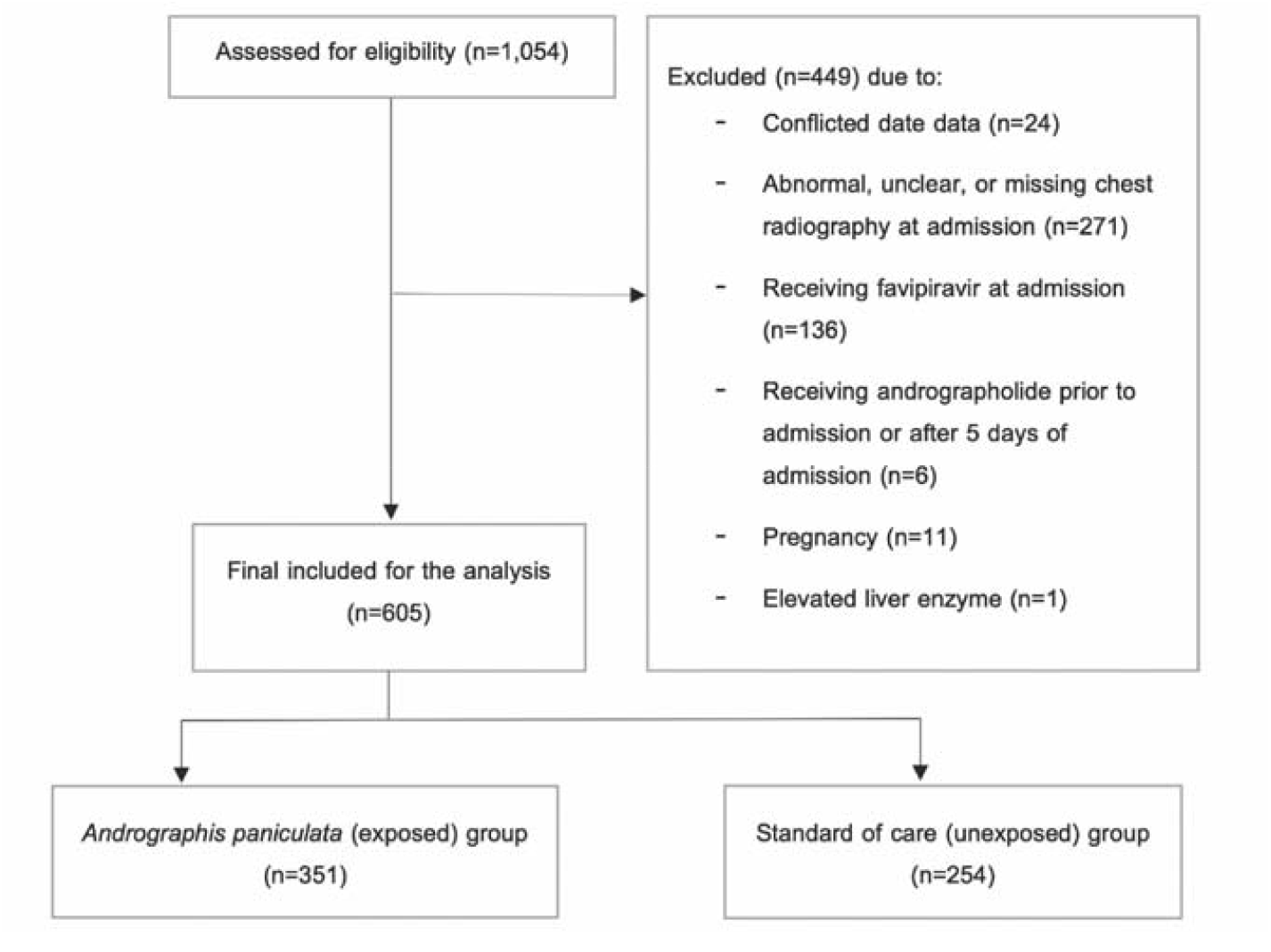
Patient flow diagram

**Table 1.** Baseline characteristics of the study populations

| Baseline characteristics | AP group<br>(n=351) | Standard of<br>care group<br>(n=254) | Total<br>(n=605) | p-value |
| --- | --- | --- | --- | --- |
| Male | 172 (49.0) | 133 (52.4) | 305 (50.4) | 0.42<sup>a</sup> |
| Age (years) | 34.84 ± 11.56 | 36.19 ± 12.13 | 35.41 ± 11.81 | 0.17<sup>b</sup> |
| Body mass index (kg/m<sup>2</sup>)<sup>†</sup> | 24.75 ± 5.08 | 23.62 ± 5.27 | 24.2 ± 5.17 | 0.32<sup>b</sup> |
| Comorbidity |  |  |  |  |
| Hypertension | 24 (6.9) | 20 (7.9) | 44 (7.3) | 0.63<sup>a</sup> |
| Diabetes | 8 (2.3) | 5 (2.0) | 13 (2.2) | 0.80<sup>a</sup> |
| Cardiovascular disease | 4 (1.1) | 1 (0.4) | 5 (0.8) | 0.41<sup>c</sup> |
| Current medications |  |  |  |  |
| ACEIs/ARBs | 14 (4.0) | 9 (3.5) | 23 (3.8) | 0.78<sup>a</sup> |
| Statins | 9 (2.6) | 7 (2.8) | 16 (2.6) | 0.88<sup>a</sup> |
| Antiplatelets | 2 (0.6) | 3 (1.2) | 5 (0.8) | 0.65<sup>c</sup> |
| Laboratory parameters<sup>†</sup> |  |  |  |  |
| WBC (10<sup>3</sup>/mm<sup>3</sup>) | 6.33 ± 2.16 | 6.43 ± 1.96 | 6.38 ± 2.05 | 0.75<sup>b</sup> |
| Lymphocyte (%) | 33.01 ± 10.22 | 30.25 ± 10.42 | 31.57 ± 10.39 | 0.09<sup>b</sup> |
| Neutrophil (%) | 56.64 ± 11.45 | 58.82 ± 11.10 | 57.77 ± 11.29 | 0.21<sup>b</sup> |
| Platelet (10<sup>3</sup>/mm<sup>3</sup>) | 228.78 ± 69.02 | 221.62 ± 70.46 | 225.07 ± 69.65 | 0.51<sup>b</sup> |
| BUN (mg/dL) | 10.79 ± 3.20 | 11.58 ± 3.84 | 11.21 ± 3.56 | 0.15<sup>b</sup> |
| Scr (mg/dL) | 0.84 ± 0.22 | 0.82 ± 0.18 | 0.83 ± 0.20 | 0.50<sup>b</sup> |
| eGFR (mL/min/1.73 m<sup>2</sup>) | 101.38 ± 18.85 | 102.12 ± 17.29 | 101.77 ± 17.99 | 0.79<sup>b</sup> |
| LDH (units/L), median<br>(IQR) | 197 (156, 231) | 192 (164, 226) | 192.5 (158, 230) | 0.69<sup>d</sup> |
| AST (units/L), median<br>(IQR) | 26 (20, 37.5) | 25 (19, 35) | 26 (20, 36) | 0.44<sup>d</sup> |
| ALT (units/L), median<br>(IQR) | 34 (22.5, 50.5) | 35 (23, 52) | 34 (23, 51) | 0.92<sup>d</sup> |
| ALP (units/L), median<br>(IQR) | 77.5 (63, 88) | 66 (58, 77) | 70 (60, 83) | 0.004<sup>d</sup> |

**Notes:** Figures represent the mean ± SD and frequency (%) unless specified elsewhere, <sup>a</sup>Chi-squared test, <sup>b</sup>Student's t test with equal variance, <sup>c</sup>Fisher's exact test, <sup>d</sup>Wilcoxon rank-sum test, <sup>†</sup>Missing values of each covariate were as follows: 83.6% (BMI), 72.4% (WBCs), 72.4% (Lymphocyte), 72.4% (Neutrophil), 72.6% (Platelet), 71.9% (BUN), 71.9% (Scr), 72.1% (eGFR), 79.2% (LDH), 72.7% (AST), 72.7% (ALT), and 72.7% (ALP), **Abbreviations:** AP; *Andrographis paniculata*, SD; standard deviation, ACEIs/ARBs; angiotensin-converting enzyme inhibitors/angiotensin II receptor blockers, BUN; blood urea nitrogen, Scr; serum creatinine, eGFR; estimated glomerular filtration rate, LDH; lactate dehydrogenase, AST; aspartate transaminase, ALT; alanine transaminase, ALP; alkaline phosphatase, WBC; white blood cell

During a median follow-up time of 7 days (IQR 6, 9 days) and a median hospital stay of 8 days (IQR 6, 10 days), 59 out of 605 participants (9.8%) developed pneumonia – an overall incidence rate of 13.35 (95% CI 10.34, 17.23) per 1,000 person-days. No deaths occurred during the study period. Comparing between groups, 37 out of 351 individuals (10.5%) in the AP group developed pneumonia, whereas 22 out of 254 patients (8.7%) in the standard of care group developed pneumonia. This corresponded to a slightly higher (but not statistically significant) incidence rate of pneumonia in the AP group (13.93 [95% CI 10.09, 19.23] per 1,000 person-days) than in the standard of care group (12.47 [95% CI 8.21, 18.94] per 1,000 person-days) (log-rank *p*-value = 0.69, Table S3 and Figure S2). According to Table S3-S4, it is worth noting that, regardless of exposure group, 1) the incidence rate of pneumonia before seven days of follow-up was higher than that afterwards, and 2) the incidence rate of pneumonia among patients aged over 60 years was drastically higher than that among younger individuals.

According to Table 2, compared to a standard of care, receiving AP was associated with increased but not statistically significant odds of having pneumonia: odds ratios (ORs) of 1.24 (95% CI 0.71, 2.16), 1.42 (95% CI 0.80, 2.54), and 1.42 (95% CI 0.79, 2.55) in an unadjusted, age-adjusted, and fully adjusted model, respectively. Furthermore, considering follow-up time and censoring yielded slightly attenuated but consistent results: hazard ratios of 1.11 (95% CI 0.66, 1.89), 1.26 (95% CI 0.74, 2.15), and 1.26 (95% CI 0.74, 2.17) in the unadjusted, age-adjusted, and fully adjusted models, respectively. Additionally, receiving AP was also associated with a slight but not significant increase in the odds of worsening symptoms. Further adjusting for BMI did not change the direction of the association (Table S6).

**Table 2.** AP use and clinical outcomes in mild COVID-19 patients

| Outcomes | Events (%) |  | Effect size (95% CI)*, *p*-value (n=605) |  |  |
| --- | --- | --- | --- | --- | --- |
|  | AP<br>(n=351) | Standard of<br>care (n=254) | Unadjusted<br>model | Age-adjusted<br>model | Fully<br>adjusted<br>model<sup>†</sup> |
| **Primary outcome: pneumonia** |  |  |  |  |  |
| Odds ratio | 37 (10.5) | 22 (8.7) | 1.24<br>(0.71, 2.16),<br>0.44 | 1.42<br>(0.80, 2.54),<br>0.23 | 1.42<br>(0.79, 2.55),<br>0.24 |
| Hazard ratio<sup>‡</sup> | 13.93<sup>§</sup><br>(10.09, 19.23) | 12.47<sup>§</sup><br>(8.21, 18.94) | 1.11<br>(0.66, 1.89)<sup>‡</sup>,<br>0.69 | 1.26<br>(0.74, 2.15)<sup>‡</sup>,<br>0.39 | 1.26<br>(0.74, 2.17)<sup>‡</sup>,<br>0.40 |
| **Secondary outcome: worsening symptoms<sup>¶</sup>** |  |  |  |  |  |
| Odds ratio | 59 (16.8) | 39 (15.4) | 1.11<br>(0.72, 1.73),<br>0.63 | 1.23<br>(0.78, 1.94),<br>0.38 | 1.22<br>(0.77, 1.94),<br>0.39 |

**Notes:** \*Effect size of outcome in the AP group, compared to the standard of care group, <sup>†</sup>Adjusting for age, diabetes, hypertension, receiving statins, and receiving ACEIs/ARBs, <sup>§</sup>Incidence rate of pneumonia per 1,000 person-days (95% confidence interval), <sup>‡</sup>Analysis using a Cox's proportional hazards model in which the fully adjusted model was additionally stratified by diabetes, <sup>¶</sup>Worsening symptoms were the composite of receiving antiviral drugs, systemic corticosteroids, or ventilator support; having oxygen saturation drop along with worsening signs and symptoms; or presenting regressive chest X-ray findings (i.e., category three or above). **Abbreviations:** AP; *Andrographis paniculata*, CI; confidence interval

Interestingly, excluding participants admitted before the 1^st^ of July 2021 (most were from the standard of care group) further strengthened the association of receiving AP with the increased odds of having outcomes. The ORs of having pneumonia in an unadjusted, age-adjusted, fully adjusted model and a model additionally adjusted for BMI were 1.83 (95% CI 0.93, 3.61), 1.94 (95% CI 0.97, 3.92), 1.88 (95% CI 0.92, 3.81), and 1.72 (95% CI 0.78, 3.79), respectively (Table S6). Also, removing previously vaccinated patients produced similar results to the main findings (Table S7).

The results from subgroup analyses are shown in Figure 2. It can be observed that sex was not an effect modifier of the association between receiving AP and pneumonia. However, the association seems stronger among the elderly (i.e., >60 years). Although AP might be related to the increased risk of pneumonia in overall populations and all *p*-values for interaction >0.05, we found the opposite direction of the associations among individuals with hypertension, receiving ACEIs/ARBs, and receiving statins.

**Figure 2.**
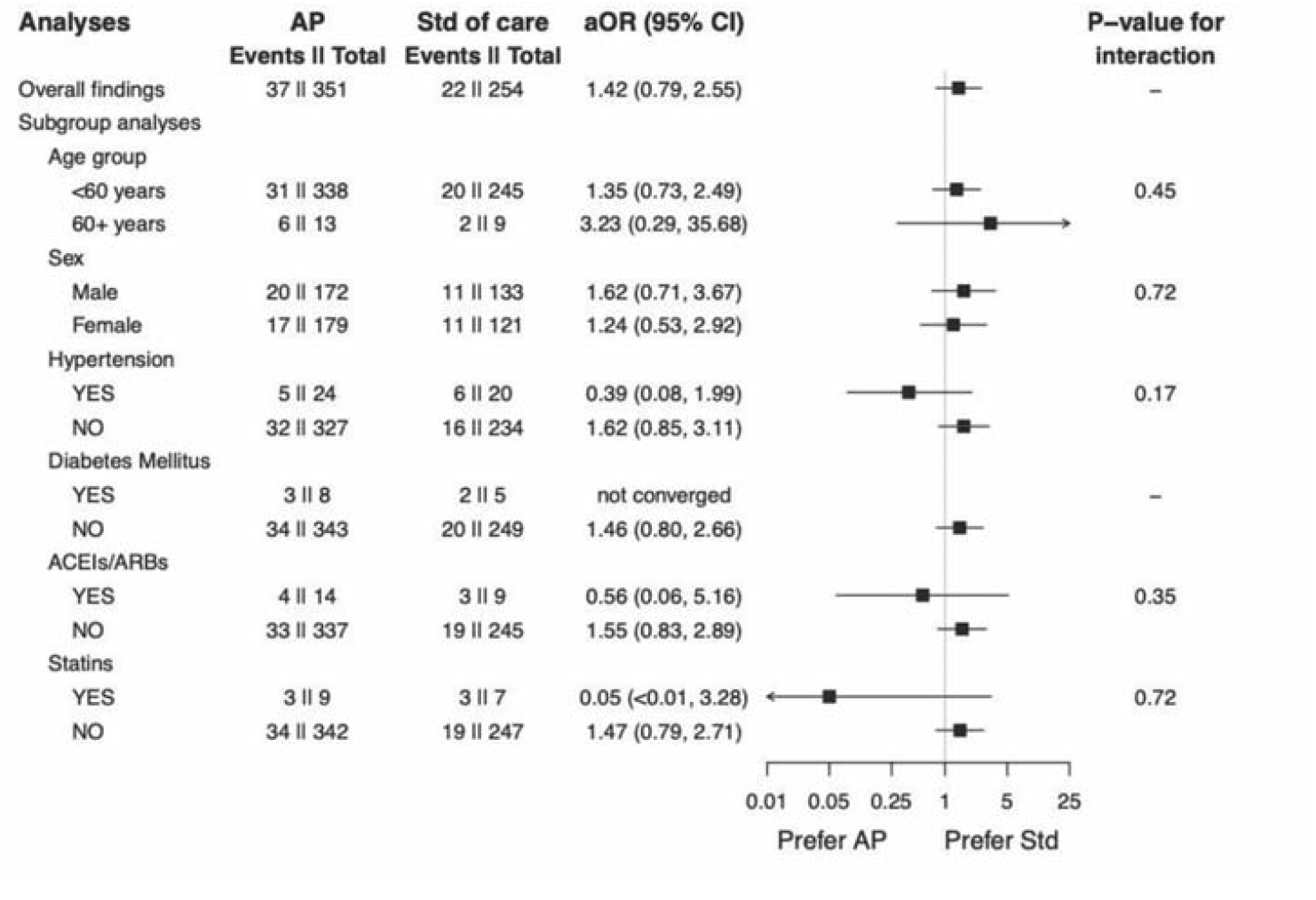
Subgroup analysis of *Andrographis paniculata* and the occurrence of pneumonia

## Discussion

### Summary of the main findings

In this retrospective cohort study of 605 hospitalised COVID-19 patients who had normal chest radiography at the time of admission, 9.8% of them developed pneumonia after a median follow-up time of 7 days. However, we did not observe an association between the use of AP and a decreased risk of pneumonia or worsening clinical symptoms. Interestingly, individuals, mainly the elderly, receiving AP were associated with an increased, but not statistically significant, risk of pneumonia and worsening clinical symptoms. Moreover, all sensitivity analyses provided consistent findings, ensuring the robustness of the main results.

### Comparison *with previous studies*

To date, clinical evidence of using AP to treat COVID-19 is still lacking. After performing a systematic search on three databases (i.e., PubMed, Google Scholar, and Thai Clinical Trial Registry), we found only two complete trials (9,10) and five ongoing trials relevant to this subject, with the largest trial of 736 patients expected to end in October 2022 (Table S2). One trial investigated the efficacy of AP in improving clinical symptoms and duration of disease in 62 mild COVID-19 patients.(9) All COVID-19 symptoms in the AP group had disappeared by day 7 (i.e., two days after completing an AP course). Compared with our observation, the median length of hospital stays before being discharged alive in the AP group and the standard of care group was eight days (IQR 6, 10 days) and seven days (IQR 6, 9 days), respectively. Therefore, the course of the disease in our study was comparable to the previous one. Another trial reported the incidence of pneumonia in the AP group (0%) and the placebo group (10.7%) after five days of treatment.(10) The figure was similar to the incidence of pneumonia in our study’s standard of care group (8.7%), confirming the validity of our collected data. Furthermore, we found that increased age, having hypertension and diabetes, and receiving ACEIs/ARBs and statins were associated with increased odds of pneumonia (Table S5). This is consistent with previous reports (15,16) and can further ensure the validity of the data used in our analyses.

In contrast to the results from a trial of Xiyanping from which andrographolide was given as an intravenous route and significant recovery was found in an active group,(11) our results were from oral administration of AP. Although there is no direct comparison study of the efficacy of AP in different dosage forms, it has been shown that andrographolide has a poor oral bioavailability (<3%) primarily due to undergoing rapid metabolism at duodenal and jejunal cells.(17)

### Strengths and limitations

To the best of our knowledge, this is the first cohort study of AP’s use in treating mild COVID-19. Admittedly, Thailand was confronted with favipiravir and vaccine shortages at the beginning of the second wave of the pandemic crisis, leading to the unproven AP’s use for this condition. Consequently, a pharmacovigilance study is required since real-world data from using AP are already available so that its efficacy and safety can be clinically ensured. Additionally, since all patients in this study were hospitalised, treatment compliance and actual consumption of AP and supportive treatment can be assured.

However, there are some limitations worth noting. First, we cannot avoid residual confounders embedded in an observational design. For instance, smoking status and mental disorders (e.g., depression) were suggested to be risk factors for developing severe COVID-19,(15,16) and this can be prevalent in people in their 30s and 40s. In addition, patients receiving AP may have a higher risk of developing pneumonia than those who do not (i.e., confounding by indication). Therefore, the observed association might result from residual confounders. However, baseline characteristics between groups were mostly similar. Furthermore, since our study populations were relatively young, many chronic conditions that can increase the risk of severe COVID-19 were rare and should not be major concerns. Additionally, the results were less likely to be confounded by favipiravir as the proportions of patients receiving favipiravir during admission were similar between groups (i.e., 9.7% in the standard of care group versus 10.6% in the AP group).

Second, our results still suffered from being underpowered despite the fact that we had analysed the data from all eligible patients by the time we conducted the research. With a sample size of 605, we had only 11% power to detect the difference in the incidence of pneumonia between the exposed (10.5%) and unexposed groups (8.7%). A total of 9,000 participants would be required to achieve at least 80% power to detect such a slight difference. However, when one carefully examines the effect sizes and the corresponding unbalanced confidence intervals (e.g., OR 1.42 [95% CI 0.79, 2.55]), increasing the sample size is prone to strengthen the harmful signal (i.e., OR or HR of more than the value of one).

Third, due to limitations of using retrospective medical records, we could not investigate the association of AP with COVID-19 symptoms, such as fatigue, cough, sputum production, anorexia, sore throat, and nasal congestion. Also, the association between AP use and CRP levels cannot be examined in our study. Although a previous trial showed that AP can reduce symptoms of mild COVID-19, open-label design and multiplicity were the major issues that could undermine the validity of the findings.(9)

In addition, we did not examine the association between the use of AP and CRP levels. Even though a previous trial showed a significant reduction of CRP levels in the AP group (*p*-value = 0.023), compared to placebo,(10) and a recent case report of CRP apheresis showed successful outcomes in seven severe COVID-19 patients,(18) further RCTs are needed before concluding the impact of AP on CRP and its role as a therapeutic target in COVID-19.

Last, data on viral strains were lacking, which might affect the external validity of our study. Nonetheless, since the incidence of pneumonia in the standard of care group in our study was similar to that in a previous trial (10) and no deaths occurred, it can be assumed that the viral strains in our study were comparable to those in the previous trial. Admittedly, the generalisability of our findings may be limited to unvaccinated patients. However, since the efficacy of the COVID-19 vaccine in reducing the severity of symptoms and pneumonia has been proven and widely accepted,(19) the role of AP in COVID-19 may, unfortunately, become less prominent over time.

### Implications

For the clinical implications, while waiting for the results from ongoing trials (Table S2) together with improved availability of favipiravir and the COVID-19 vaccine, we suggested that physicians should suspend the use of AP to treat COVID-19. This is because we observed potentially harmful signal without proof of benefit, even if causality cannot be established. For the research implications, a multicentre collaboration is required to achieve a sufficient sample size and confirm our findings. In addition, the safety parameters of using AP were rarely monitored. We noticed that less than one-fourth of patients receiving AP underwent liver and renal function tests at baseline and were rarely measured afterwards. Although a previous study has shown the safety of AP used in other indications,(20) the safety of using such a high dose of AP in COVID-19 is still unclear and needs further investigation.

## Conclusions

In summary, we had insufficient evidence to show the association of the use of AP for the treatment of mild COVID-19 with a decreased risk of pneumonia. The results from ongoing randomised controlled trials should provide insight into this issue. In the meantime, using AP in this condition should be cautious or suspended.

## Data Availability

All data produced in the present study are available upon reasonable request to the corresponding authors.

## Abbreviations

ACEIs: angiotensin-converting enzyme inhibitors
ALP: alkaline phosphatase
ALT: alanine transaminase
aOR: adjusted odds ratio
AP: *Andrographis paniculata*
ARBs: angiotensin II receptor blockers
AST: aspartate transaminase
BMI: body mass index
BUN: blood urea nitrogen
CI: confidence interval
COVID-19: coronavirus disease 2019
CRP: c-reactive protein
CXR: chest-X rays
DAGs: directed acyclic graphs
eGFR: estimated glomerular filtration rate
HR: hazard ratio
ICD-10: the 10^th^ revision of the International Classification of Diseases
IQR: interquartile range
LDH: lactate dehydrogenase
MAR: missing at random
MICE: multiple imputation by chained equations
OR: odds ratio
SARS-CoV-2: severe acute respiratory syndrome coronavirus 2
Scr: serum creatinine
SD: standard deviation
Std of care: standard of care
STROBE: Strengthening the Reporting of Observational Studies in Epidemiology
T2DM: type 2 diabetes mellitus
RT–PCR: Real-time polymerase chain reaction
WBC: white blood cell

## Data availability statement

The original contributions presented in the study are included in the article/supplementary material, further inquiries can be directed to the corresponding authors.

## Ethics statement

The report of this study followed the Strengthening the Reporting of Observational Studies in Epidemiology (STROBE) guidance for reporting cohort studies. The ethical committee for clinical research of Phrae Hospital approved this study (no. 70/2564).

## Authors’ contribution

JT, SK, and NN conceptualised the study objectives and designed and collected the data. JT, SK, and NN contributed to the literature review, data cleaning, data analyses, and interpretation of the findings. JT prepared an initial manuscript, and NN further developed subsequent manuscripts. SK, PT, NP, LS, BB, and CS critically revised the initial manuscript, and all authors participated in further revisions. The final manuscript was read and approved by all authors before submission.

## Acknowledgements

We thank members of the staff at Phrae Hospital for facilitating the data collection process of this study.

