## Supplementary Appendix for "Use of *Andrographis paniculata* (Burm.f.) Wall. ex Nees and risk of pneumonia in hospitalised patients with mild COVID-19: a retrospective cohort study"

**Table S1** STROBE checklist for cohort study

|  | Item No. | Recommendation | Page No. |
| --- | --- | --- | --- |
| Title and abstract | 1 | (a) Indicate the study’s design with a commonly used term in the title or the abstract  (b) Provide in the abstract an informative and balanced summary of what was done and what was found | 1, 3 |
| Introduction | | | |
| Background/rationale | 2 | Explain the scientific background and rationale for the investigation being reported | 5 |
| Objectives | 3 | State specific objectives, including any prespecified hypotheses | 5 |
| Methods | | | |
| Study design | 4 | Present key elements of study design early in the paper | 5 |
| Setting | 5 | Describe the setting, locations, and relevant dates, including periods of recruitment, exposure, follow-up, and data collection | 5-6 |
| Participants | 6 | (a) Give the eligibility criteria, and the sources and methods of selection of participants. Describe methods of follow-up  (b) For matched studies, give matching criteria and number of exposed and unexposed | 5-6 |
| Variables | 7 | Clearly, define all outcomes, exposures, predictors, potential confounders, and effect modifiers. Give diagnostic criteria, if applicable | 6 |
| Data sources/ measurement | 8* | For each variable of interest, give sources of data and details of methods of assessment (measurement). Describe comparability of assessment methods if there is more than one group | 6 |
| Bias | 9 | Describe any efforts to address potential sources of bias | 6-7 |
| Study size | 10 | Explain how the study size was arrived at | 6-7 |
| Quantitative variables | 11 | Explain how quantitative variables were handled in the analyses. If applicable, describe which groupings were chosen and why | 6-7 |
| Statistical methods | 12 | (a) Describe all statistical methods, including those used to control for confounding  (b) Describe any methods used to examine subgroups and interactions  (c) Explain how missing data were addressed  (d) If applicable, explain how loss to follow-up was addressed  (e) Describe any sensitivity analyses | 6-7 |
| Results | | | |
| Participants | 13* | (a) Report numbers of individuals at each stage of study—eg numbers potentially eligible, examined for eligibility, confirmed eligible, included in the study, completing follow-up, and analysed  (b) Give reasons for non-participation at each stage  (c) Consider use of a flow diagram | 7,  Fig 1 |
| Outcome data | 15* | Report numbers of outcome events or summary measures over time | 7-8, Table S3-4, Fig S2 |
| Main results | 16 | (a) Give unadjusted estimates and, if applicable, confounder-adjusted estimates and their precision (eg, 95% confidence interval). Make clear which confounders were adjusted for and why they were included  (b) Report category boundaries when continuous variables were categorized  (c) If relevant, consider translating estimates of relative risk into absolute risk for a meaningful time period | 7-8, Table 2 |
| Other analyses | 17 | Report other analyses done—eg analyses of subgroups and interactions, and sensitivity analyses | Fig 2, Table S6-7 |
| Discussion | | | |
| Key results | 18 | Summarise key results with reference to study objectives | 8 |
| Limitations | 19 | Discuss limitations of the study, taking into account sources of potential bias or imprecision. Discuss both direction and magnitude of any potential bias | 9-10 |
| Interpretation | 20 | Give a cautious overall interpretation of results considering objectives, limitations, multiplicity of analyses, results from similar studies, and other relevant evidence | 8-10 |
| Generalisability | 21 | Discuss the generalisability (external validity) of the study results | 10 |
| Other information | | | |
| Funding | 22 | Give the source of funding and the role of the funders for the present study and, if applicable, for the original study on which the present article is based | 10 |

*Give such information separately for cases and controls in case-control studies, and, if applicable, for exposed and unexposed groups in cohort and cross-sectional studies.

### Quality assurance of *Andrographis paniculata* product (exposure) used in Phrae Hospital

Certificate of analysis**:** The analysis and quality control reported from the Medicinal Plant Research Institute and the regional Medical Sciences Center 1/1, Chiang Rai, Thailand

| **Test** | **Results** | **Methods** | **Acceptance criteria** |
| --- | --- | --- | --- |
| Total lactones content, calculated as andrographolide | 6.0% w/w | Thai Herbal Pharmacopoeia | Not less than 6.0% w/w |
| Andrographolide content | 4.16% w/w | Thai Herbal Pharmacopoeia | Not less than 1.0% w/w of andrographolide |
| Basic chemical tests  (Phytochemical tests) | Complied with the standard | Thai Herbal Pharmacopoeia 2017 | Positive |
| Chemical Identification by Thin-Layer Chromatography | Complied with the standard | Thai Herbal Pharmacopoeia 2017 | Complied with the standard of *Andrographis paniculata* (Burm. f.) Wall. ex Nees |
| Moisture content analysis by gravimetric analysis | 5.5% w/w | Thai Herbal Pharmacopoeia 2017 | Not more than 11.0% w/w |
| Acid-insoluble ash | 0.2% w/w | Thai Herbal Pharmacopoeia 2017 | Not more than 2.0% w/w |
| Ethanol (85%)-soluble extractive | 17.9% w/w | Thai Herbal Pharmacopoeia 2017 | Not less than 13.0% w/w |
| Water-soluble extractive | 20.9% w/w | Thai Herbal Pharmacopoeia 2017 | Not less than 18.0% w/w |
| Weight variation | Complied with the standard | Thai Herbal Pharmacopoeia 2017 | Not more than 2 capsules with weight variation beyond the range of +/- 10% and no individual capsule with weight variation beyond the range of +/- 20% |
| Disintegration time | 8 minutes | Thai Pharmacopoeia 1997 Volume II Part 1 | All shall be disintegrated within 30 minutes. |
| Total aerobic microbial count per gram or milliliter | Less than 10 | Thai Herbal Pharmacopoeia | Not more than 500,000 cfu per gram or milliliter |
| Total combined yeast and mold count per gram or milliliter | Less than 10 | Thai Herbal Pharmacopoeia | Not more than 50,000 cfu per gram or milliliter |
| Bile-tolerant gram-negative bacteria per gram or milliliter | Less than 10 | Thai Herbal Pharmacopoeia | Not more than 1,000 cfu per gram or milliliter |
| *Salmonella* spp. per 10 grams or milliliter | Not found | Thai Herbal Pharmacopoeia | Not found per 10 grams or milliliter |
| *Escherichia coli* per gram or milliliter | Not found | Thai Herbal Pharmacopoeia | Not found per grams or milliliter |
| *Clostridium* spp. per gram or milliliter | Not found | Thai Herbal Pharmacopoeia | Not found per grams or milliliter |
| Identification A, B, C | A=Purplish color  B=Yellow color  C=Find dark violet spot chromatogram | Thai Herbal Pharmacopoeia | Complied with the standard |
| Loss on drying | 5.61% w/w | Thai Herbal Pharmacopoeia | Not more than 11.0% w/w |
| Ethanol-soluble extractive | 18.49% w/w | Thai Herbal Pharmacopoeia | Not less than 13.0% w/w |
| Water-soluble extractive | 21.31% w/w | Thai Herbal Pharmacopoeia | Not less than 18.0% w/w |
| Acid-insoluble ash | 0.004% w/w | Thai Herbal Pharmacopoeia | Not more than 2.0% w/w |

### Abbreviations: cfu; colony-forming unit, % w/w; percentage weight by weight

### Modified Rama Co‐RADS for first chest X-rays in confirmed COVID-19 patients

Source(1)

Categories 1‐6 and C are for the initial chest X-rays (CXR) in a new patient or the first CXR in a home isolation or community isolation patient. In this regard, whenever the patient has an old CXR before having COVID‐19, the newly performed CXR after confirmed COVID‐19 should be interpreted using categories 1, 2, 3, 4, 5, 6, or C. Details of each category were as follows:

- Category 1: Normal chest X‐ray or no abnormality detected
- Category 2: Presence of minor abnormalities unrelated to COVID‐19 (e.g., mild cardiomegaly, aortic atherosclerosis, scoliosis, old fractures)
- Category C: Low likelihood or atypical for COVID‐19 pneumonia, but with other clinically significant diseases (e.g., bacterial pneumonia, active TB, CHF, pneumothorax, pleural effusion, malignancy) unrelated to COVID‐19
- Category 3: Equivocal/indeterminate opacities, which may be due to acute or residual/post‐COVID‐19 pneumonia or pseudolesions
- Category 4: Single or multifocal poorly defined ground‐glass opacities or consolidations in one lung, suspicious for early/mild acute or post‐COVID‐19 pneumonia ± fibrosis‐like changes
- Category 5: Multifocal, peripheral, poorly defined ground‐glass opacities or consolidations with or without rounded morphology involving any zones of both lungs, typical for moderate/severe acute or post‐COVID‐19 pneumonia ± fibrosis‐like changes
- Category 6: Acute or post‐COVID‐19 pneumonia with its related conditions or complications (e.g., atelectasis, PE, pulmonary infarction, OP, AFOP, secondary infection, pneumothorax, pneumomediastinum)

### Systematic search

#### PubMed

("COVID-19"[All Fields] OR "COVID-19"[MeSH Terms] OR "COVID-19 Vaccines"[All Fields] OR "COVID-19 Vaccines"[MeSH Terms] OR "COVID-19 serotherapy"[All Fields] OR "COVID-19 serotherapy"[Supplementary Concept] OR "covid 19 nucleic acid testing"[All Fields] OR "covid 19 nucleic acid testing"[MeSH Terms] OR "covid 19 serological testing"[All Fields] OR "covid 19 serological testing"[MeSH Terms] OR "covid 19 testing"[All Fields] OR "covid 19 testing"[MeSH Terms] OR "sars cov 2"[All Fields] OR "sars cov 2"[MeSH Terms] OR "Severe Acute Respiratory Syndrome Coronavirus 2"[All Fields] OR "NCOV"[All Fields] OR "2019 NCOV"[All Fields] OR (("coronavirus"[MeSH Terms] OR "coronavirus"[All Fields] OR "COV"[All Fields]) AND 2019/11/01:3000/12/31[Date - Publication])) AND ("andrographolide"[Supplementary Concept] OR "Andrographis paniculata"[Text Word] OR "Andrographis paniculata extract"[Text Word] OR "andrographolide"[Text Word])

Searching on 15/12/2021 without language restriction yielded 43 results and found no relevant clinical studies.

#### Google Scholar

("andrographis" OR "Andrographis paniculata" OR "andrographolide") AND ("sars-cov2" OR "sars cov 2" OR "covid-19" OR "covid") restricted to the range between 2019 and 2021

Searching on 15/12/2021 without language restriction yielded 1,460 results: 2 relevant studies.(2,3)

#### Thai Clinical Trial Registry (TCTR)

There are seven currently registered RCTs. Two are completed RCTs: one small pilot study with no published results (n=6), and another one can be found at <https://doi.org/10.1101/2021.07.08.21259912> (n=57).(3) Additionally, there are five ongoing RCTs (Table S2), including the largest one (n=736) anticipated to be completed in October 2022.(4)

**Table S2** Summary of five ongoing trials of *Andrographis paniculata* used in COVID-19 patients in Thailand

| **Trial** | **TCTR 20210514003**(5) | **TCTR 20210609001**(6) | **TCTR 20210809004**(7) | **TCTR 20210906002**(8) | **TCTR 20211022002**(4) |
| --- | --- | --- | --- | --- | --- |
| Main sponsor | DTAM | Chulabhorn Royal Academy | Thammasat university | Chulalongkorn University | Health Systems Research Institute |
| Patient | Asymptomatic COVID-19 patients (n=160) | Mild to moderate COVID-19 patients (n=146) | Asymptomatic or mild COVID-19 patients (n=186) | Mild to moderate COVID-19 patients (n=160) | Asymptomatic or mild COVID-19 patients without pneumonia (n=736) |
| Intervention | Andrographolide 180 mg/day for 5 days | Andrographolide 180 mg/day for 5 days + Favipiravir 3.6 g day 1 then 1.6 g/day for 4 days | Andrographolide 180 mg/day | Andrographolide 180 mg for 5 days + Favipiravir (200 mg) 9x2 for 1 day then 4x2 for 4 days. | Andrographis capsule 180 mg/day for 5 days |
| Comparator | Placebo | Favipiravir 3.6 g day 1 then 1.6 g/day * 4 days | Placebo | Favipiravir (200 mg) 9x2 for 1 day then 4x2 for 4 days alone. | Placebo |
| Primary outcome | Hospitalisation rate during 14 days of follow-up | Clinical stable or improvement of symptom at day 4 | Symptoms and severity score until discharge | Proportion of patients developing severe pneumonia at 6 months | Pneumonia at day 10 |
| Study design | Randomised, double blind, placebo-controlled trial | Randomised, double blind, placebo-controlled trial | Randomised, double blind, placebo-controlled trial | Randomised, open label, active-controlled trial | Randomised, double blind, placebo-controlled trial |
| Status | Pending (ethic submitted) | Completed | Completed | Pending (ethic submitted) | Recruiting |
| Updated in TCTR | 20 Oct 2021 | 5 Apr 2022 | 13 Mar 2022 | 6 Sep 2021 | 2 Mar 2022 |

**Notes:** Updated from TCTR on 2 Jul 2022, Abbreviations: DTAM; Department of Thai Traditional and Alternative Medicine, TCTR; Thai Clinical Trial Registry

**Table S3** Incidence rate of pneumonia by follow-up time

| **Group** | **Events** | **Person-days of**  **follow-up** | **Incidence rate**  **per 1,000 person-days (95% CI)** |
| --- | --- | --- | --- |
| ***Andrographis paniculata* group** | | | |
| Before 7 days | 31 | 2,203 | 14.07 (9.90, 20.01) |
| After 7 days | 6 | 453 | 13.25 (5.95, 29.48) |
| Overall | 37 | 2,656 | 13.93 (10.09, 19.23) |
| **Standard of care group** | | | |
| Before 7 days | 20 | 1,470 | 13.61 (8.78, 21.09) |
| After 7 days | 2 | 294 | 6.80 (1.70, 27.20) |
| Overall | 22 | 1,764 | 12.47 (8.21, 18.94) |
| **Total** | | | |
| Before 7 days | 51 | 3,673 | 13.89 (10.55, 18.27) |
| After 7 days | 8 | 747 | 10.71 (5.36, 21.41) |
| Overall | 59 | 4,420 | 13.35 (10.34, 17.23) |

**Table S4** Incidence rate of pneumonia according to age groups

| **Group** | **Events** | **Person-days of**  **follow-up** | **Incidence rate**  **per 1,000 person-days (95% CI)** |
| --- | --- | --- | --- |
| ***Andrographis paniculata* group** | | | |
| Age <60 | 31 | 2,557 | 12.12 (8.53, 17.24) |
| Age 60+ | 6 | 99 | 60.61 (27.23, 134.90) |
| Overall | 37 | 2,656 | 13.93 (10.09, 19.23) |
| **Standard of care group** | | | |
| Age <60 | 20 | 1,683 | 11.88 (7.67, 18.42) |
| Age 60+ | 2 | 81 | 24.69 (6.18, 98.73) |
| Overall | 22 | 1,764 | 12.47 (8.21, 18.94) |
| **Total** | | | |
| Age <60 | 51 | 4,240 | 12.03 (9.14, 15.83) |
| Age 60+ | 8 | 180 | 44.44 (22.23, 88.87) |
| Overall | 59 | 4,420 | 13.35 (10.34, 17.23) |

**Table S5** The association between baseline characteristics and incident pneumonia

| Baseline characteristics | Pneumonia | | Total  (n=605) | *p*-value |
| --- | --- | --- | --- | --- |
|  | Yes (n=59) | No (n=546) |  |  |
| Male | 31 (52.5) | 274 (50.2) | 305 (50.4) | 0.73 |
| Age (years) | 44.71 (12.57) | 34.40 (11.29) | 35.41 (11.81) | <0.001 |
| Body mass index (kg/m^2^)^†^ | 26.04 (4.07) | 23.96 (5.28) | 24.2 (5.17) | 0.23 |
| Comorbidity | | | | |
| Hypertension | 11 (18.6) | 33 (6.0) | 44 (7.3) | <0.001 |
| Diabetes | 5 (8.47) | 8 (1.5) | 13 (2.2) | <0.001 |
| Cardiovascular disease | 0 | 5 (0.9) | 5 (0.8) | 1.00 |
| Current medications | | | | |
| ACEIs/ARBs | 7 (11.9) | 16 (2.9) | 23 (3.8) | 0.001 |
| Statins | 6 (10.2) | 10 (1.8) | 16 (2.6) | <0.001 |
| Antiplatelets | 0 | 5 (0.9) | 5 (0.8) | 1.00 |
| Laboratory parameters^†^ | | | | |
| WBC (10^3^/mm^3^) | 6.31 (1.70) | 6.40 (2.16) | 6.38 (2.05) | 0.80 |
| Lymphocyte (%) | 26.79 (9.36) | 33.03 (10.28) | 31.57 (10.39) | 0.001 |
| Neutrophil (%) | 62.69 (11.11) | 56.27 (10.95) | 57.77 (11.29) | 0.002 |
| Platelet (10^3^/mm^3^)  Mean (SD)  Median (IQR) | 218.74 (70.12)  209 (155, 255) | 227.01 (69.67)  218 (181, 261) | 225.07 (69.65)  215.5 (178, 260) | 0.52 |
| BUN (mg/dL) | 11.23 (3.85) | 11.20 (3.49) | 11.21 (3.56) | 0.96 |
| Scr (mg/dL) | 0.84 (0.17) | 0.83 (0.21) | 0.83 (0.20) | 0.89 |
| eGFR (mL/min/1.73m^2^) | 97.52 (15.16) | 103.05 (18.62) | 101.77 (18.0) | 0.09 |
| LDH (units/L), median (IQR) | 188.5 (155, 228) | 201.5 (172.5, 230.5) | 192.5 (158, 230) | 0.14 |
| AST (units/L), median (IQR) | 26 (22, 38) | 26 (20, 36) | 26 (20, 36) | 0.25 |
| ALT (units/L), median (IQR) | 35 (25, 47) | 34 (22, 55) | 34 (23, 51) | 0.81 |
| ALP (units/L), median (IQR) | 70 (58, 88) | 70.5 (61, 83) | 70 (60, 83) | 0.96 |

**Table S6** Sensitivity analyses of *Andrographis paniculata* and clinical outcomes

| Outcomes | Odds ratio (95% confidence interval)*, *p*-value | | | |
| --- | --- | --- | --- | --- |
|  | Unadjusted  model | Age-adjusted model | Fully adjusted model^†^ | Additional adjusting model^††^ |
| Total participants (n=605, events=59) | | | | |
| Primary outcome | | | | |
| Pneumonia | 1.24 (0.71, 2.16),  0.44 | 1.42 (0.80, 2.54),  0.23 | 1.42 (0.79, 2.55),  0.24 | 1.29 (0.68, 2.46),  0.44 |
| Pneumonia^‡^ | 1.12 (0.66, 1.89)^‡^,  0.68 | 1.27 (0.75, 2.16)^‡^,  0.38 | 1.27 (0.74, 2.17)^‡^,  0.39 | 1.20 (0.67, 2.13)^‡^,  0.54 |
| Secondary outcome | | | | |
| Worsening symptoms¶ | 1.11 (0.72, 1.73),  0.63 | 1.23 (0.78, 1.94),  0.38 | 1.22 (0.77, 1.94),  0.39 | 1.21 (0.74, 1.98),  0.45 |
| Excluding participants admitted before the 1^st^ of July, 2021 (n=545, events=49) | | | | |
| Primary outcome | | | | |
| Pneumonia | 1.83 (0.93, 3.61),  0.08 | 1.94 (0.97, 3.92),  0.06 | 1.88 (0.92, 3.81),  0.08 | 1.72 (0.78, 3.79),  0.18 |
| Pneumonia^‡^ | 1.57 (0.82, 3.02)^‡^,  0.18 | 1.57 (0.82, 3.03)^‡^,  0.17 | 1.55 (0.80, 3.02)^‡^,  0.19 | 1.48 (0.72, 3.03)^‡^,  0.29 |
| Secondary outcome | | | | |
| Worsening symptoms¶ | 1.34 (0.82, 2.21),  0.25 | 1.39 (0.83, 2.33),  0.21 | 1.38 (0.82, 2.33),  0.22 | 1.32 (0.74, 2.36),  0.19 |

**Notes**: *Analysis using multiple imputation by chain equation (MICE), ^†^Adjusting for age, diabetes, hypertension, receiving statins, and ACEIs/ARBs, ^§^Incidence rate of pneumonia per 1,000 person-days (95% confidence interval), ^‡^Cox’s proportional hazards model, in which fully adjusted model was additionally stratified by diabetes, and effect sizes were reported as hazard ratio (95% CI). ^††^Additional adjusting model was a fully adjusting model with further adjusting for body mass index. ^¶^Worsening symptoms were the composite of receiving antiviral drugs, systemic corticosteroids, or ventilator support; having oxygen saturation drop along with worsening signs and symptoms; or presenting regressive chest X-ray findings (i.e., category three or above).

**Table S7** Sensitivity analysis of AP use and clinical outcomes in mild COVID-19 patients, excluding those who had previous COVID-19 vaccination at least 1 shot

| **Outcomes** | **Events (%)** | | **Effect size (95% CI)*, *p*-value (n=603)** | | |
| --- | --- | --- | --- | --- | --- |
|  | **AP**  **(n=350)** | **Standard of care (n=253)** | **Unadjusted model** | **Age-adjusted model** | **Fully adjusted model^†^** |
| **Primary outcome: pneumonia** | | | | | |
| Odds ratio | 36 (10.3) | 22 (8.7) | 1.20  (0.69, 2.10), 0.51 | 1.37  (0.77, 2.45),  0.29 | 1.37  (0.76, 2.46),  0.30 |
| Hazard ratio^‡^ | 13.56^§^  (9.78, 18.80) | 12.56^§^  (8.27, 19.08) | 1.07  (0.63, 1.83)^‡^, 0.79 | 1.21  (0.71, 2.06)^‡^, 0.48 | 1.21  (0.70, 2.08)^‡^, 0.50 |
| **Secondary outcome: worsening symptoms**^¶^ | | | | | |
| Odds ratio | 58 (16.6) | 39 (15.4) | 1.09  (0.70, 1.70),  0.70 | 1.20  (0.76, 1.89),  0.44 | 1.19  (0.75, 1.89),  0.45 |

**Notes:** *Effect size of outcome in the AP group, compared to the standard of care group, ^†^Adjusting for age, diabetes, hypertension, receiving statins, and receiving ACEIs/ARBs, ^§^Incidence rate of pneumonia per 1,000 person-days (95% confidence interval), ^‡^Analysis using a Cox’s proportional hazards model in which the fully adjusted model was additionally stratified by diabetes, ^¶^Worsening symptoms were the composite of receiving antiviral drugs, systemic corticosteroids, or ventilator support; having oxygen saturation drop along with worsening signs and symptoms; or presenting regressive chest X-ray findings (i.e., category three or above). **Abbreviations**: AP; *Andrographis paniculata*, CI; confidence interval

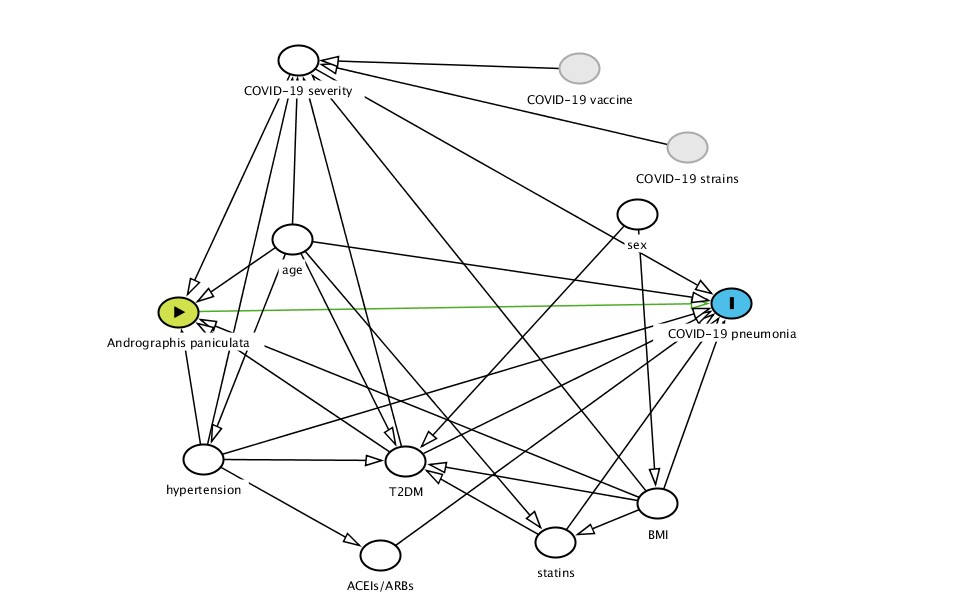

**Figure S1** Directed acyclic graphs (DAGs) of the association between *Andrographis paniculata* and the incidence of pneumonia

**Abbreviations**: ACEIs/ARBs; angiotensin converting enzyme inhibitors/angiotensin receptor blockers, BMI; body mass index, T2DM; type 2 diabetes mellitus, **Sources**:(9,10)

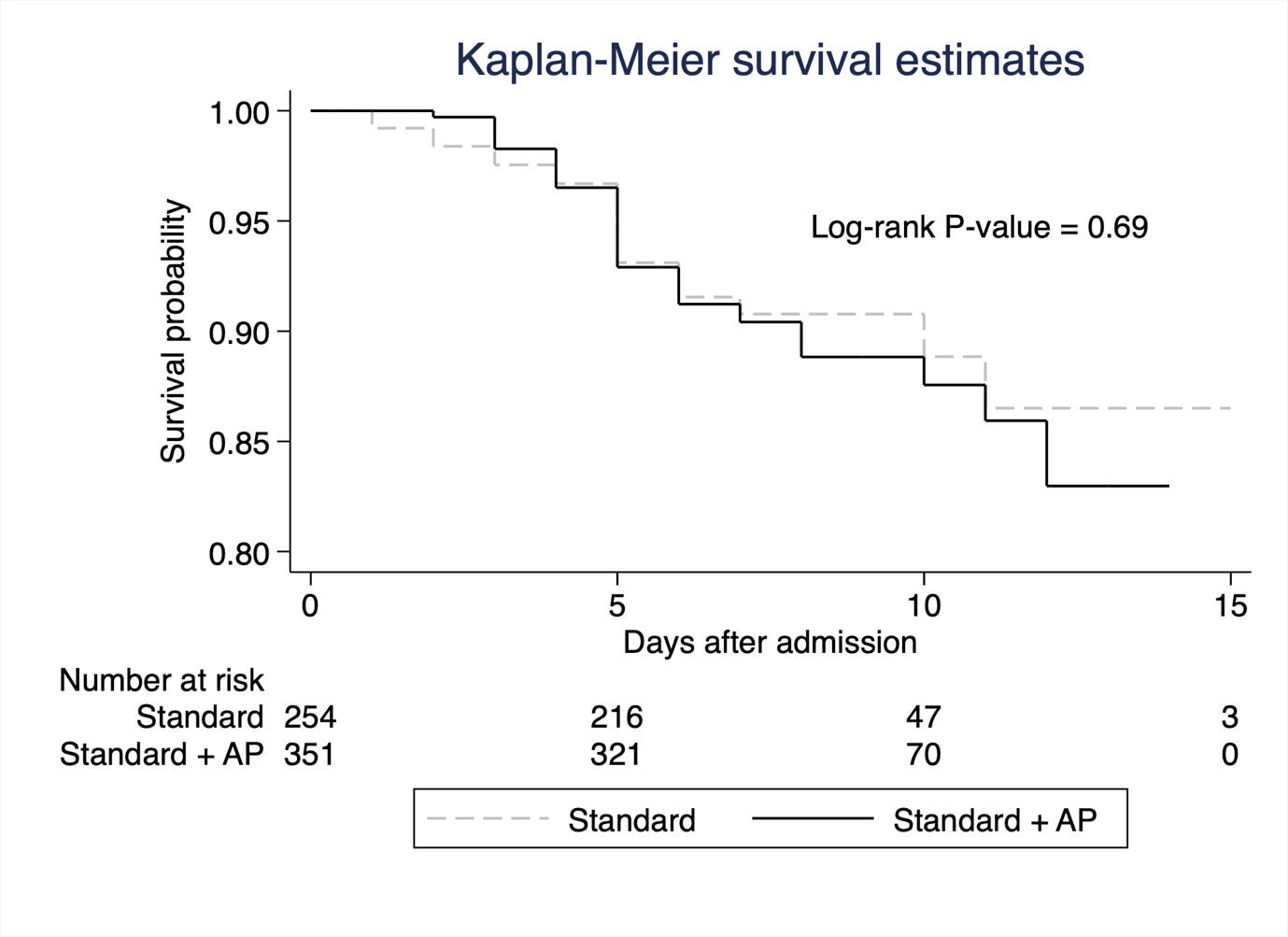

**Figure S2** Probability of a pneumonia-free event by exposure

**Abbreviation**: AP; *Andrographis paniculata*

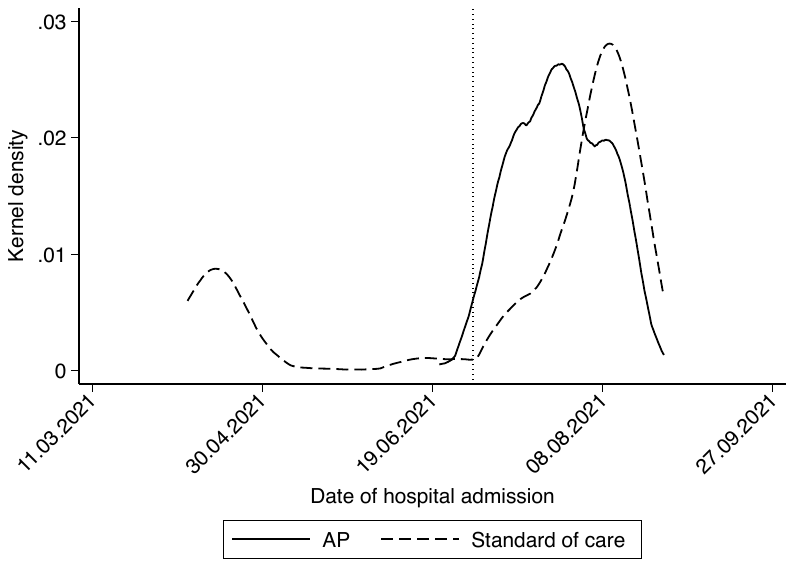

**Figure S3** The distribution of hospital admission dates between the *Andrographis paniculata* group and the standard of care group

**Notes:** A vertical dotted line represents the 1^st^ of July 2021. **Abbreviations:** AP; *Andrographis paniculata*
